# The impact of cross-reactive immunity on the emergence of SARS-CoV-2 variants

**DOI:** 10.1101/2022.09.20.22280161

**Authors:** R.N. Thompson, E. Southall, Y. Daon, F.A. Lovell-Read, S. Iwami, C.P. Thompson, U. Obolski

## Abstract

A key feature of the COVID-19 pandemic has been the emergence of SARS-CoV-2 variants with different transmission characteristics. However, when a novel variant arrives in a host population, it will not necessarily lead to many cases. Instead, it may fade out, due to stochastic effects and the level of immunity in the population. Immunity against novel SARS-CoV-2 variants may be influenced by prior exposures to related viruses, such as other SARS-CoV-2 variants and seasonal coronaviruses, and the level of cross-reactive immunity conferred by those exposures. Here, we investigate the impact of cross-reactive immunity on the emergence of SARS-CoV-2 variants in a simplified scenario in which a novel SARS-CoV-2 variant is introduced after an antigenically related virus has spread in the population. We use mathematical modelling to explore the risk that the novel variant invades the population and causes a large number of cases, as opposed to fading out. If cross-reactive immunity is complete (i.e. someone infected by the previously circulating virus is no longer susceptible to the novel variant), the novel variant must be more transmissible than the previous virus to invade the population. However, in a more realistic scenario in which cross-reactive immunity is partial, we show that it is possible for novel variants to invade, even if they are less transmissible than previously circulating viruses. This is because partial cross-reactive immunity effectively increases the pool of susceptible hosts that are available to the novel variant compared to complete cross-reactive immunity. Furthermore, if previous infection with the antigenically related virus assists the establishment of infection with the novel variant, as has been proposed following some experimental studies, then even variants with very limited transmissibility are able to invade the host population. Our results highlight that fast assessment of the level of cross-reactive immunity conferred by related viruses on novel SARS-CoV-2 variants is an essential component of novel variant risk assessments.

## 1 Introduction

When a new SARS-CoV-2 variant first arrives in a host population, a key question for policy makers is whether or not it will become widespread. For this to occur, two steps are required: introduction and invasion. First, the variant must arrive in the host population, either through de novo mutation or by importation from elsewhere (introduction). Second, the variant must then spread from person to person and cause a large number of cases, as opposed to simply fading out (invasion). Following introduction, a range of factors affect the risk that a novel variant will invade, including its inherent transmissibility and the connectivity of the location in which it is introduced [1,2]. An additional crucial factor in this process is the background level of immunity to the new variant in the host population. For example, a feature of the Omicron (B.1.1.529) variant that allowed it to become widespread is its ability to evade immunity from past infection or vaccination, at least partially, meaning that the background immunity level was low [3–6].

Mathematical modelling has often been used to explore the impact of cross-reactive immunity between pathogen strains on the dynamics of infectious disease outbreaks [7–12]. During the COVID-19 pandemic, models have provided real-time insights into the risk from novel variants. For example, Bhatia *et al*. [13] extended existing methods for estimating pathogen transmissibility [14,15] to enable the transmissibility of novel variants to be assessed, including estimating the infectiousness of the Alpha (B.1.1.7), Beta (B.1.351) and Gamma (P.1) variants relative to the wild type virus (the SARS-CoV-2 virus that first emerged in Wuhan, China). Dyson *et al*. [16] analysed epidemiological data from England, and projected the course of the outbreak in that country if a variant emerged with different transmission characteristics. They warned that a variant with high transmissibility or substantial immune escape properties had the potential to generate large numbers of infections and hospitalisations.

Meanwhile, experimental and statistical studies have explored the effects of prior infections with related viruses on infections with different SARS-CoV-2 variants. Some studies have found previous infections with other SARS-CoV-2 variants to have a protective effect. For example, Wratil *et al*. [17] demonstrated that a combination of infection and vaccination induced hybrid immunity is protective against SARS-CoV-2 variants including the Omicron variant. A recent analysis of infection data from Portugal found that previous SARS-CoV-2 infections were protective against infection with the BA.5 Omicron subvariant, with the level of protection particularly high in individuals who were previously infected by the BA.1 or BA.2 Omicron subvariants [18]. However, some studies have indicated that prior infection with other SARS-CoV-2 variants may instead have a detrimental effect on subsequent infections with novel SARS-CoV-2 variants. For example, infection with the SARS-CoV-2 wild type was found to inhibit the immune response to infections with the Omicron variant among triple vaccinated healthcare workers [6].

Similarly to the cross-reactive immunity conferred by other SARS-CoV-2 variants, the impact of prior infections with seasonal coronaviruses on subsequent infections by SARS-CoV-2 is also unclear. Some analyses have found that previous infections with seasonal coronaviruses are likely to be protective against SARS-CoV-2 infection. The SARS-CoV-2 spike protein can be divided into the S1 and S2 subunits. The S1 subunit contains an antigenically variable receptor binding domain, while the S2 subunit is more conserved between coronaviruses. Kaplonek *et al*. [19] showed that SARS-CoV-2 S2 antibody responses are associated with milder COVID-19 symptoms, suggesting that previous infection with seasonal coronaviruses may lead to COVID-19 infections being less severe. Furthermore, strong and multispecific cross-reactive T-cell responses induced by seasonal coronavirus infection prior to SARS-CoV-2 infection have been associated with protection against SARS-CoV-2 infection in seronegative healthcare workers [6,20].

In contrast, there is also evidence that previous infections with seasonal coronaviruses can have detrimental effects on susceptibility to and outcomes of infection with SARS-CoV-2. With respect to disease outcomes, McNaughton *et al*. [21] showed that prior immunity to seasonal coronaviruses was positively associated with fatal outcomes in individuals with severe COVID-19. Similar results were found by Smit *et al*. [22] in an independent cohort. Conflicting results to Kaplonek *et al*. [19] were found by Garrido *et al*. [23], who found that S2 antibody responses were associated with greater disease severity. With respect to susceptibility, Wratil *et al*. [24] demonstrated that reactive cross-immunity imparted by seasonal coronaviruses may increase susceptibility to SARS-CoV-2. Additionally, a modelling analysis by Pinotti *et al*. [25] has suggested that the general trend of increased severity of SARS-CoV-2 infections in older individuals may be explained by an increased chance that older individuals have been exposed to seasonal coronaviruses.

Given this conflicting evidence in the literature, and to help understand the possible effects of prior infections on the risk of emergence of SARS-CoV-2 variants, in this study we develop a mathematical model considering two viruses: a novel SARS-CoV-2 variant and an antigenically related virus that has previously spread in the population. We investigate the factors affecting the risk that the novel variant invades the host population. We assume that infection with the previously circulating virus affects the chance of successful infection with, and subsequent transmission of, the novel variant, considering scenarios in which prior infection is either protective (partially or completely) or detrimental. We show that the level of cross-reactive immunity between novel SARS-CoV-2 variants and antigenically related viruses is a key factor determining whether or not a novel variant will invade the host population. This highlights the need to conduct a rapid assessment of the level of cross-reactive immunity between previously circulating viruses and newly emerged SARS-CoV-2 variants whenever a novel SARS-CoV-2 variant is introduced into a new host population.

## 2 Methods

### 2.1 Epidemiological model

We consider the introduction of a novel variant to a population consisting of *N* hosts. We assume that prior immunity has been conferred by infections with a related virus that has already spread within the host population. Assuming that this previously circulating virus follows dynamics that are characterised by the standard (determinstic) SEIR model, the number of individuals in the population that have been previously infected by that virus is given by the solution, *N*_*p*_, to the final size equation [9],

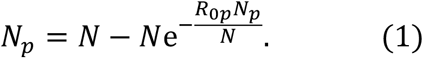

In this expression, *R*_0*p*_ is the reproduction number of the previously circulating virus, which we assume accounts for any interventions that were introduced (prior to, or immediately after, its arrival in the host population) to limit its spread. We assume that *N*_*p*_ individuals were previously infected by that virus (we round *N*_*p*_ to the nearest integer value), and the remaining *N* − *N*_*p*_ individuals in the population are immunologically naïve (i.e. they do not carry any immunity against the novel variant).

We then model the emergence of the novel variant. If an individual has previously been infected by the related virus, their susceptibility to the novel variant is assumed to be modified by a factor 1 − *α* (relative to the susceptibility of a host who has not previously been infected by the related virus). Consequently, if *α* > 0, prior infection with the related virus is protective against infection with the novel variant. If instead if *α* = 0, then earlier infection with the related virus does not affect the risk of infection with the novel variant. If *α* < 0, earlier infection with the related virus promotes infection with the novel variant. Similarly, the infectiousness of a host infected with the novel variant who has previously been infected by the related virus is modified by a factor 1 − *ε* (relative to the infectiousness of a host who has not previously been infected by the related virus). Again, positive (negative) values of *ε* reflect scenarios in which prior infection with the related virus reduces (increases) the infectiousness of an individual who is infected with the novel variant.

Transmission dynamics for the novel variant are also modelled using an SEIR model, but with two main differences compared to the dynamics of the previously circulating virus. First, the SEIR model for the novel variant is extended to account for cross-reactive immunity conferred by the related virus. Second, since we are modelling invasion, we use a stochastic model in which, in each simulation of the model, the novel variant may either invade the host population or fade out with few infections. The analogous deterministic model to the stochastic model that we consider for the novel variant is given by:

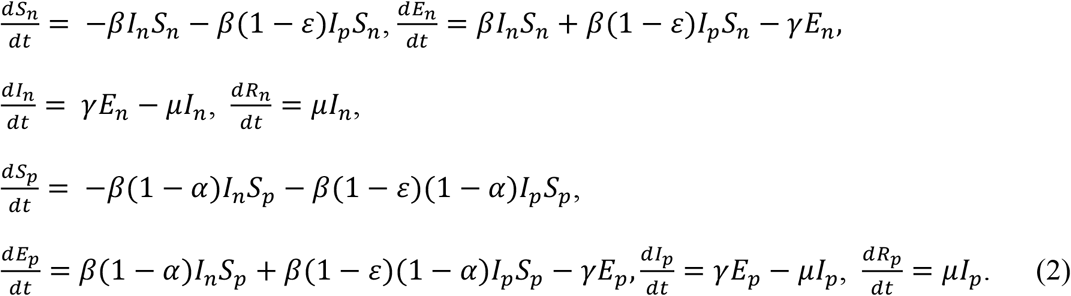

In these equations, the variables *S*_*n*_, *E*_*n*_, *I*_*n*_ and *R*_*n*_ refer to the infection status (with the novel variant) of individuals who have not been infected previously by the related virus, and *S*_*p*_, *E*_*p*_, *I*_*p*_ and *R*_*p*_ refer to the infection status of individuals who have previously been infected by the related virus. A schematic illustrating the transitions of individuals between these states, and the rates at which those transitions occur, is shown in Fig 1A. The parameter *β* is the infection rate parameter, and the mean latent period and infectious period are 1/*γ* days and 1/*µ* days, respectively. We define the reproduction number of the novel variant to be 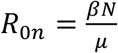, reflecting the transmission potential of the novel variant if the host population is entirely immunologically naïve.

**Figure 1.**
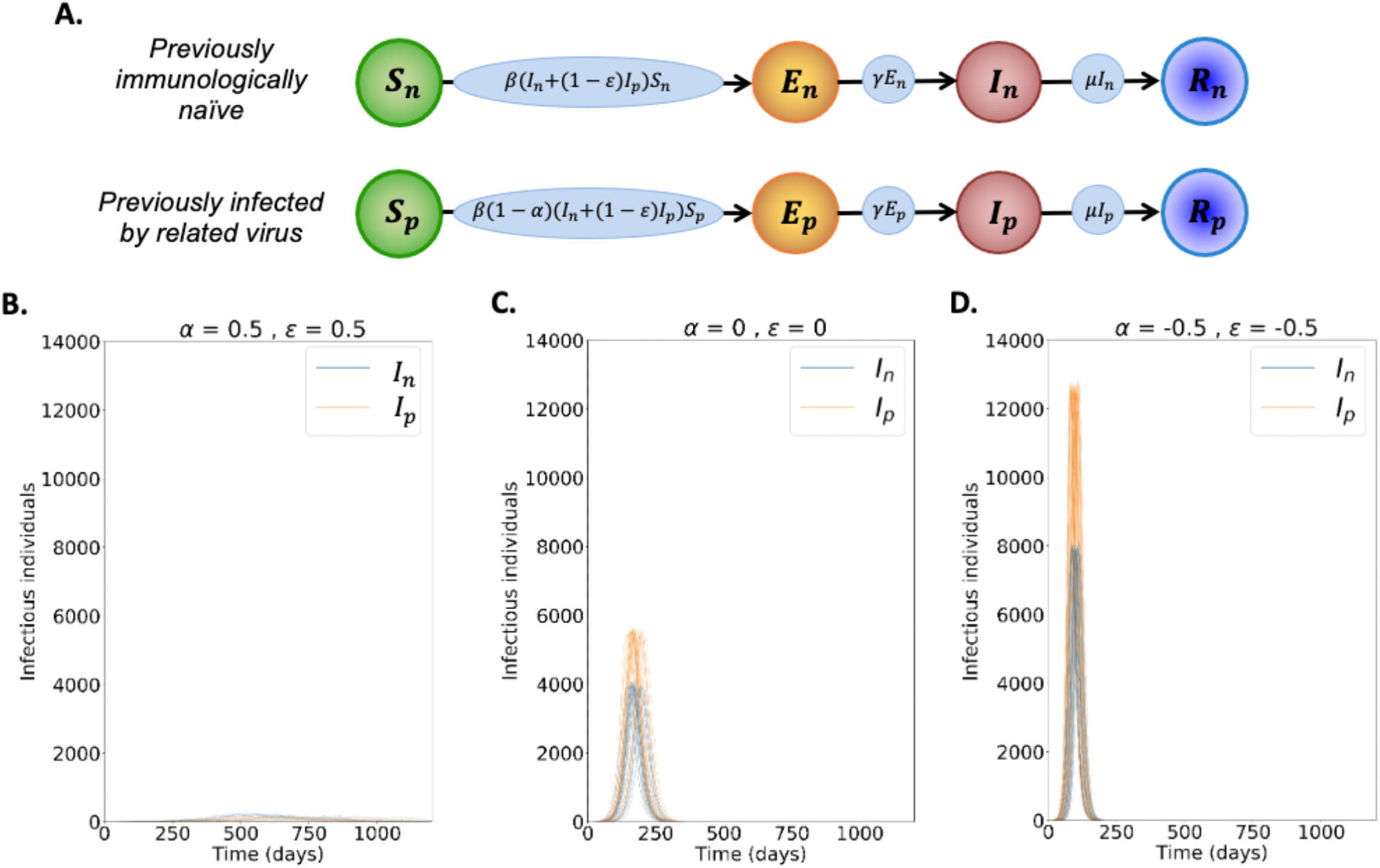
Dynamics of the novel variant invading a population in which a related virus has previously spread. A. Schematic showing the transitions (and their rates) in the stochastic model of novel variant invasion (the analogous stochastic model to system of equations (2)). B. Realisations of 50 stochastic simulations of the model, for *R*_0*n*_= 2 and with protective cross-reactive immunity (*α* = *ε* = 0.5; other parameter values are as stated in Table 1). Orange lines represent the number of individuals infected by the novel variant who were previously infected by the related virus (*I*_*p*_), and blue lines represent the number of individuals infected by the novel variant who were previously immunologically naïve (*I*_*n*_). C. Analogous to panel B, but with no cross-reactive immunity (*α* = *ε* = 0). D. Analogous to panel B, but with cross-reactive immunity instead promoting infection with the novel variant (*α* = *ε* = −0.5). Simulations were initiated with a single infected, previously immunologically naïve individual (*I*_*n*_ = 1), with all other individuals susceptible (*S*_*n*_ = *N − N*_*p*_ − 1 and *S*_*p*_ = *N*_*p*_, where *N*_*p*_ is the solution of the final size equation for the previously circulating virus, equation (1), rounded to the nearest integer value).

As noted above, since we are interested in the risk of invasion of the novel variant, we use the analogous stochastic model to system of equations (2) rather than solving the differential equations numerically. We run model simulations using the direct method version of the Gillespie stochastic simulation algorithm [26] until the novel variant fades out (i.e. *E*_*n*_ + *I*_*n*_ + *E*_*p*_ + *I*_*p*_ reaches zero). The parameter values used in our main analyses are given in Table 1.

**Table 1.** Illustrative parameter values used in model simulations.

| Parameter | Meaning | Value used |
| --- | --- | --- |
| $N$ | Size of local host population | 100,000 |
| $1/\gamma$ | Mean latent period of novel variant | 5 days [57,58] |
| $1/\mu$ | Mean infectious period of novel variant | 8 days [59–61] |
| $R_{0p}$ | Reproduction number of previously circulating virus (accounting for interventions) | 1.5, so that $N_p = 58,281$ individuals are assumed to have been infected by the previously circulating virus (~58% of the population) |
| $R_{0n}$ | Reproduction number of novel variant (accounting for interventions) | Varies (see figures) |
| $\beta$ | Transmission rate of novel variant | Set so that $R_{0n} = \frac{\beta N}{\mu}$ |
| $\alpha$ | Reduction (positive) or increase (negative) in susceptibility due to cross-reactive immunity | Varies (see figures) |
| $\varepsilon$ | Reduction (positive) or increase (negative) in infectiousness due to cross-reactive immunity | Varies (see figures) |

### 2.2 Risk of invasion

When the novel variant is introduced, we approximate the probability that it invades the population analytically. To do this, we assume that infections occur according to a branching process [27–30]. Specifically, we denote by *q*_*ij*_ the probability that the novel variant fails to invade the host population, starting from *i* currently infected individuals who are immunologically naïve and *j* currently infected individuals who were previously infected by the related virus. In this analysis, “currently infected” individuals refer to those who are either exposed or infectious, since exposed and infectious individuals are each expected to infect the same number of other hosts in future. This is because exposed individuals are not yet infectious, and only start generating infections when they move into the infectious compartments in the model.

We then consider the probability of the novel variant failing to establish in the host population starting from a single currently infected individual who was previously immunologically naïve, *q*_10_. As in similar previous branching process analyses [31–34], we consider the various possibilities for what happens next: either that individual infects another individual who was also previously immunologically naïve (with probability 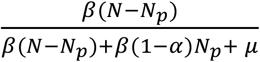); or, that individual infects someone who was previously infected with the related virus (with probability 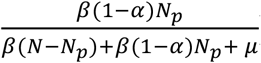); or, that individual recovers without infecting anyone else (with probability 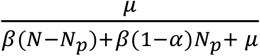). Applying the law of total probability therefore gives

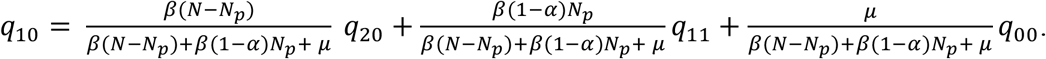

Instead starting from a single infected individual who was previously infected by the related virus gives

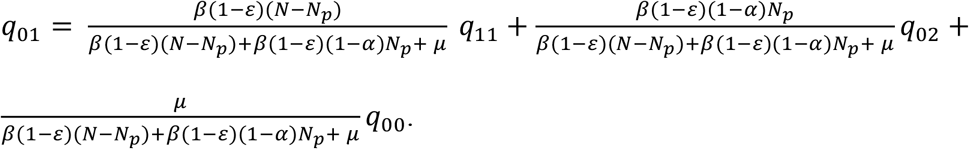

We then assume that infections occur according to a branching process (so that *q*_20_ = *q*_10_^2^; as infection lineages failing to establish starting from two infected hosts requires the infection lineages from both infected hosts to fail independently [31–33]). Making similar approximations throughout the equations above, and noting that *q*_00_ = 1 (since the novel variant will not invade if there are no infected individuals) gives

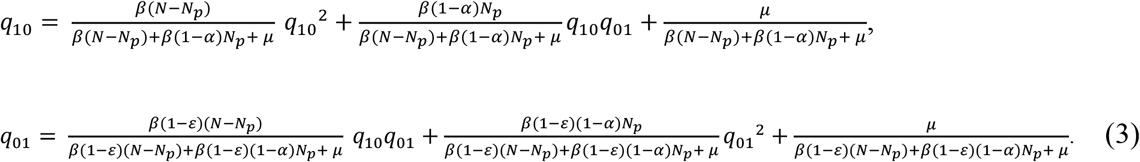

The probability of invasion starting from one infected individual who was previously immunologically naïve, *p*_10_, and the probability of invasion starting from one infected individual who was previously infected by the related virus, *p*_01_, are then given by *p*_10_ = 1 *− q*_10_ and *p*_01_ = 1 − *q*_01_, where *q*_10_ and *q*_01_ are the minimal non-negative solutions of system of equations (3) [35].

### 2.3 Special cases

In general, we solve system of equations (3) numerically. However, an analytic solution is possible in some special cases.

For example, in a scenario in which previous infection with the related virus is entirely protective against infection with the novel variant, then *α* = 1. In that case, since a previously infected individual cannot be infected with the novel variant, then *p*_01_ does not apply. However, in that scenario, 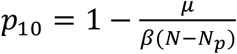 (whenever 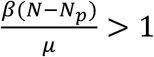; otherwise, the novel variant will never establish in the host population). This can be seen by substituting *α* = 1 into the first equation of system of equations (3), solving the resulting quadratic equation for *q*_10_ (taking the minimal non-negative solution [35]), and then calculating *p*_10_ = 1 − *q*_10_. In a scenario in which the related virus has not previously spread in the host population, then this solution for *p*_10_ is identical to the classic branching process estimate for the probability of a major outbreak, 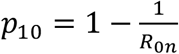 [28,36,37].

Alternatively, we can consider a scenario in which prior infection with the related virus eliminates the infectiousness of a host infected by the novel variant (i.e. the individual can become infected, but the virus cannot then establish, so onwards transmission cannot occur). In that case, *ε* = 1 and so – in a similar fashion to above – we obtain 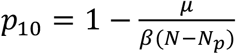 (whenever 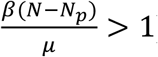 and *p*_01_ = 0.

Again, in a scenario in which a related virus has not previously spread in the population, this is the classic estimate for the probability of a major outbreak, 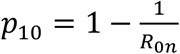 [28,36,37].

Finally, in a scenario in which previous infection by the related virus does not affect the dynamics of the novel variant (so that *α* = *ε* = 0), we expect the risk of novel variant invasion to be independent of whether or not the initial infected individual has previously been infected by the related virus. In other words, we expect *q*_10_ = *q*_01_ In this case, system of equations (3) reduces to a single quadratic equation for *q*_10_. Taking the minimal non-negative solution of that equation [35] indicates that 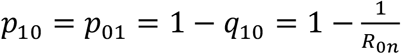 (whenever *R*_0*n*)_ > 1; otherwise the novel variant will never establish in the host population).

## 3 Results

To investigate the effects of prior infection by an antigenically related virus on the epidemiological dynamics of a newly emerged variant, we first ran stochastic simulations of the analogous stochastic model to system of equations (2). Representative time series of the dynamics illustrate that, if the novel variant successfully spreads in the host population, outbreaks tend to have a lower peak number of infections and have a longer duration when cross-reactive immunity has a protective effect (Fig 1B), compared to when prior infection by the related virus has no effect (Fig 1C). In contrast, if prior infection by the related virus instead promotes infection by the novel variant, outbreaks tend to have a higher peak number of infections and a shorter duration (Fig 1D).

However, rather than focusing on the dynamics of outbreaks once the novel variant has invaded the host population, our main goal was to quantify the risk of the novel variant successfully invading in the first place (as opposed to fading out with few cases). We therefore calculated the risk of the novel variant invading the population, starting from the introduction of a single case to the population (Fig 2). We not only calculated this quantity by numerically solving system of equations (3) (Fig 2 - red dotted and dashed lines), but also confirmed that these numerical approximations matched estimates of the invasion probability obtained using large numbers of simulations of the stochastic model (Fig 2 - black dots and crosses).

**Figure 2.**
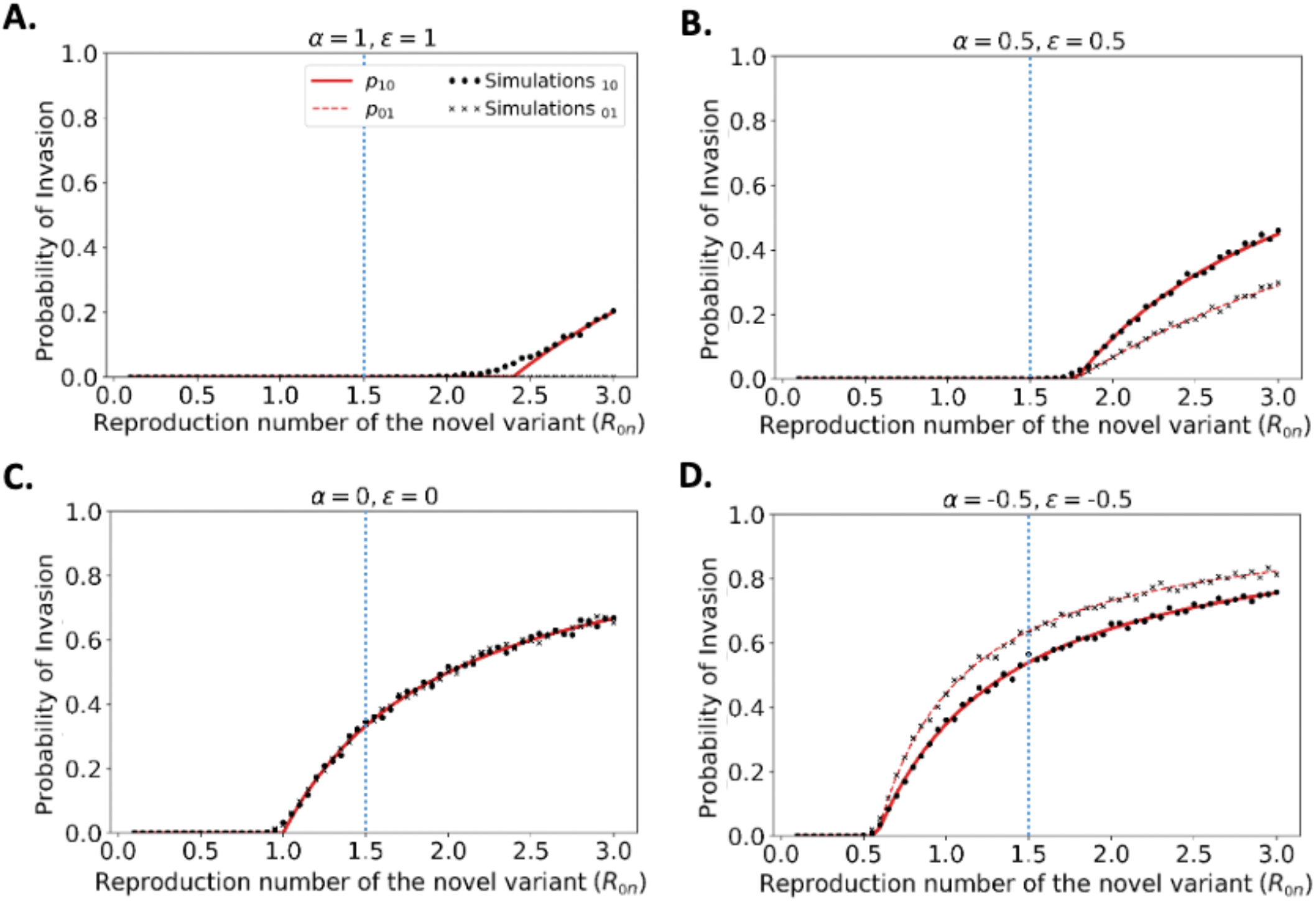
Probability of the novel variant invading the host population, starting from the introduction of a single infectious individual. A. The probability of the novel variant invading under an assumption of perfectly protective cross-reactive immunity (*α* = *ε* = 1). Results are shown both for analytic approximations of the invasion probability calculated using system of equations (3) (either starting from a single infected individual who was previously immunologically naïve (red solid) or starting from a single infected individual who was previously infected by the related virus (red dashed)) and for the invasion probability calculated using stochastic simulations (either starting from a single infected individual who was previously immunologically naïve (black dots) or starting from a single infected individual who was previously infected by the related virus (black crosses)). The vertical blue dotted line represents the reproduction number of the previously circulating virus (*R*_0*p*_ = 1.5). B. Analogous results to panel A, but with partial protective cross-reactive immunity (*α* = *ε* = 0.5). C. Analogous results to panel A, but with no cross-reactive immunity (*α* = *ε* = 0). D. Analogous to panel A, but with cross-reactive immunity instead promoting infection with the novel variant (*α* = *ε* = −0.5). In the simulations, the probability of invasion was calculated as the proportion of simulations in which the number of simultaneously infected individuals (*I*_*n*_ + *I*_*p*_) exceeded 15 at any time. As in Fig 1, the division of the host population between individuals who were previously immunologically naïve and those who were infected by the related virus was calculated based on the final size equation for the previously circulating virus (equation (1)). Other parameter values used are shown in Table 1.

We found that, when previous infection with the related virus is completely protective against the novel variant (i.e. *α* = *ε* = 1), then the reproduction number of the novel variant must be higher than the reproduction number of the antigenically related virus in order for the novel variant to invade. Specifically, in Fig 2A (in which *R*_0*p*_ = 1.5, as marked by the vertical blue dotted line), the probability of the novel variant invading the host population is zero unless *R*_0*n*_ > *R*_0*p*_, and indeed remains zero whenever *R*_0*n*_ is only slightly larger than *R*_0*p*_. This can be explained analytically as follows. The previously circulating virus will spread around the population until sufficiently many individuals have been infected for herd immunity (to the previous virus) to be reached. This occurs when 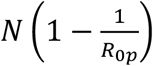 individuals have become infected [38]. However, at this point, infections do not stop immediately: there is an “overshoot” in infections while transmission slows and the previously circulating virus fades out. As a result, a lower bound on the final size of the outbreak caused by the previously circulating virus is 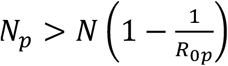. As noted in the Methods (Special cases), in a scenario involving complete cross-reactive immunity, the novel variant can only invade the population if 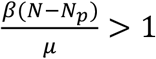, or equivalently 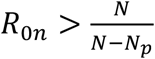. Substituting the lower bound for *N*_*p*_ into this expression shows that invasion of the novel variant requires *R*_0*n*_ > *R*_0*p*_.

In contrast, if cross-reactive immunity is only partial, then the novel variant may invade for lower values of _0*p*_ (Fig 2B). This can include scenarios in which *R*_0*n*_ < *R*_0*p*_ (in some cases lying between those shown in Fig 2B and Fig 2C). As noted in the Methods, when previous infection by the antigenically related virus does not affect the epidemiological dynamics of the novel variant, then the novel variant can only invade if *R*_0*n*_ > 1 (Fig 2C), mirroring the classical result for models in which cross-reactive immunity is not accounted for [36]. Finally, in scenarios in which prior infection by the related virus promotes infection with the novel variant, the novel variant can invade even if *R*_0*n*_ is small. This includes scenarios in which *R*_0*n*_ < 1 (Fig 2D).

In Fig 2, we note that the immune status of the initial infected individual affects the risk that the novel variant will invade the host population. In particular, when cross-reactive immunity is protective, we found that the probability of invasion is higher if the initially infected host had not previously been infected by the related virus (Fig 2B). In contrast, if cross-reactive immunity promotes infection with the novel variant, then the probability of the novel variant invading is higher if the initial infection arose in an individual who had previously been infected with the related virus (Fig 2D).

We then explored how the probability of invasion of the novel variant depends on the susceptibility- and infectiousness-modifying effects of cross-reactive immunity individually (Fig 3). We found that the values of *α* and *ε* affect the probability of a major outbreak differently. This is because, starting from a single infected individual, the number of infections generated by that individual is crucial in determining whether or not a novel variant will invade. If the first infected host infects multiple others, then all of those individuals’ transmission lineages must fade out in order for invasion to fail to occur. Hence, invasion is much more likely to occur if the first infected individual infects many individuals. Starting from a single infected individual who was not previously infected by the related virus, only susceptibility-reducing immunity (characterised by *α*) affects the number of infections generated by the first infected individual. As such, the probability of invasion in this case is more sensitive to *α* than to *ε* (Fig 3A).

**Figure 3.**
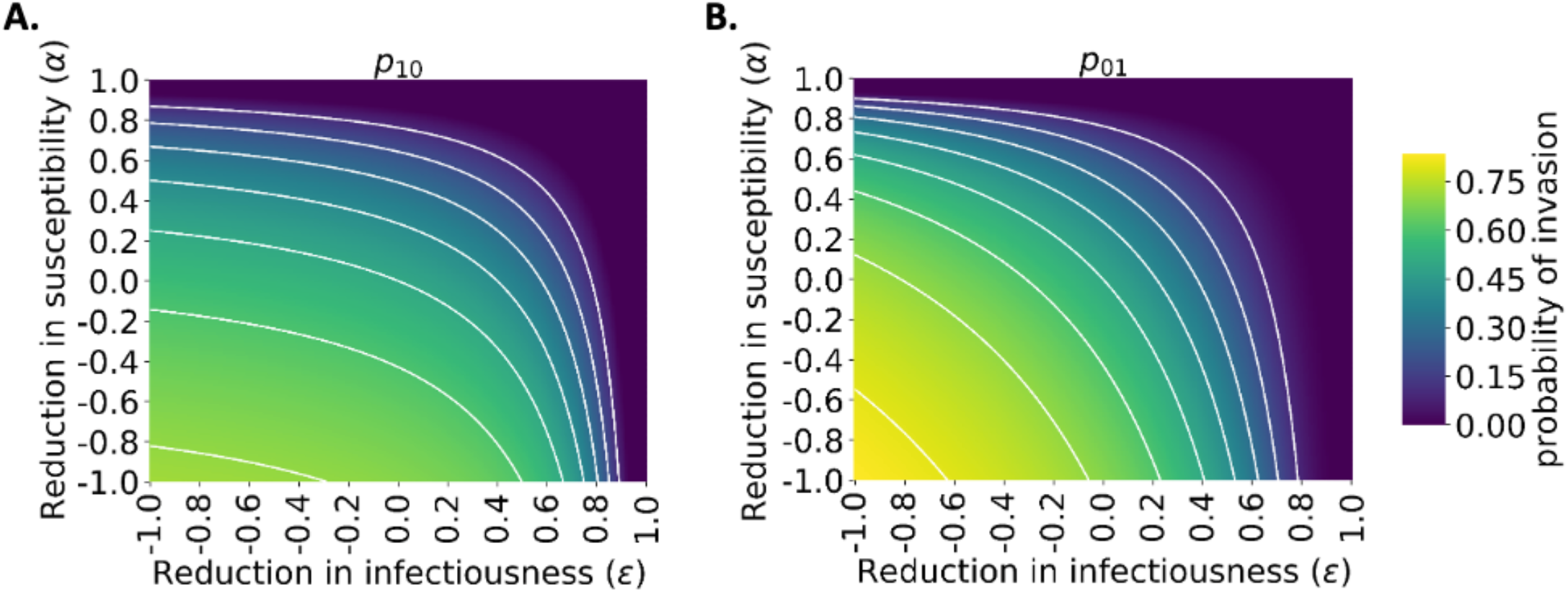
Probability of the novel variant invading the host population, starting from the introduction of a single infectious individual, for different levels of cross-reactive immunity affecting susceptibility and infectiousness individually. The invasion probability is approximated analytically using system of equations (3). A. The probability of the novel variant invading, starting from a single infected individual who was previously immunologically naïve. B. Analogous to panel A, but starting from a single infected individual who was previously infected by the related virus. White lines represent contours of constant probability of invasion of the novel variant. As in Fig 1, the division of the host population between individuals who were previously immunologically naïve and those who were infected by the related virus was calculated based on the final size equation for the previously circulating virus (equation (1)). Other parameter values used are shown in Table 1.

In contrast, if the first infected individual was previously infected by the related virus, then infectiousness-reducing immunity (characterised by *ε*) also affects the probability of this individual infecting any other member of the population. In fact, *ε* then affects all potential transmissions generated by the first infected individual, whereas *α* only affects potential transmissions to part of the population (those individuals who were previously infected by the related virus). In that scenario, the probability of a major outbreak is therefore slightly more sensitive to *ε* than *α* (Fig 3B). The different effects of susceptibility-reducing and infectiousness-reducing cross-reactive immunity therefore explain the asymmetric nature of the contours about the diagonal *ε* = *α* in Fig 3.

## 4 Discussion

The epidemiological dynamics of the COVID-19 pandemic have been shaped by the emergence of different SARS-CoV-2 variants. However, not all variants that have appeared have propagated and caused a large number of cases. Most novel variants have faded out, with relatively few variants being responsible for the vast majority of SARS-CoV-2 infections.

Here, we have developed a mathematical model to investigate the impact of cross-reactive immunity (generated by previous infections by related viruses) on the probability that a newly introduced variant will invade the host population. We found that, if prior infection with a related virus has a strong protective effect, then the novel variant must be more infectious than the related virus to be able to invade the host population (Fig 2A). If instead, however, the previously circulating virus has a very weak protective effect or no protective effect on infection with the novel variant, then the risk of invasion of the novel variant is unaffected by the outbreak of the related virus, and so the invasion probability matches the well-known estimate for the “probability of a major outbreak” (Fig 2C) [28,36,37]. If prior infection with the related virus promotes infection by the novel variant, as has been proposed in some studies exploring the impact of prior infections by SARS-CoV-2 or seasonal coronaviruses on infections with SARS-CoV-2 variants, then even novel variants with limited transmissibility are able to invade (Fig 2D).

We further showed that the immune status of the first individual in the population infected by the novel variant affects the probability that the novel variant invades (Figs 2 and 3). This is in turn influenced by the pathway by which the novel variant is introduced into the host population. If the variant is introduced from elsewhere, for example by an incoming traveller [1,39], then it may be introduced by someone who was not previously infected by the related virus. If instead it appears as a result of mutation from a related virus within the local population (as was likely the case for the emergence of the Alpha variant in Kent, England [40]), then the initial infected case would be an individual who was previously infected by the related virus.

Previous modelling studies have explored the risk of a novel virus invading when it is introduced to a host population, including scenarios in which the pathogen evolves to facilitate emergence [41–45]. Of significant relevance to our study, Hartfield and Alizon [46] applied a branching process model to investigate the invasion probability in a scenario in which a resident pathogen strain that confers cross-immunity is spreading in the host population, and considered Chikungunya virus as a case study. Those authors demonstrated that a simple estimate for the probability of a major outbreak overestimates the invasion probability in that scenario, due to the potential for depletion of susceptible individuals by the resident strain over the timescale of invasion of the novel virus. Echoing this result in a single strain setting, Sachak-Patwa *et al*. [47] showed that simple estimates for the probability of a major outbreak are overestimates if the pathogen enters the host population during a vaccination campaign, again due to susceptible depletion occurring within the period of the pathogen either invading or fading out. Other researchers have investigated the emergence of a novel pathogen strain that is introduced to the population when a resident strain is endemic [48]. In contrast to previous studies, we focus on a scenario in which a related virus has already spread widely around the host population and caused a completed outbreak, rather than being resident in the host population. An additional novel aspect of the current study is that we conduct a thorough investigation of the effect of different levels of cross-reactive immunity, including scenarios in which prior infection with an antigenically related virus can promote infection with the novel variant. Although such scenarios may seem counterintuitive, recent evidence suggests that there is a clear possibility that infection-promoting cross-reactivity may occur, as described in the Introduction.

To understand general principles governing the relationship between cross-reactive immunity and the risk of invasion of a novel variant, we constructed the simplest possible model in this study. Further developments could involve including additional epidemiological and evolutionary detail in our transmission model, particularly if it is to be used to predict emergence of specific variants rather than to understand general principles. For example, in the model considered here, the infectious period of individuals infected by the novel variant is assumed to follow an exponential distribution. However, gamma distributions have been found to represent observed epidemiological periods more accurately than exponential distributions [49,50], and gamma-distributed infectious periods can be incorporated into calculations of invasion probabilities [51,52]. We also assumed a fixed level of cross-reactive immunity for all individuals who were previously infected by the related virus. In reality, immunity is heterogeneous between different previously infected hosts, and is likely to wane over time [53,54]. Waning immunity has been included previously in a range of epidemiological models [55,56], and is a target for future study in the modelling framework presented here.

In summary, understanding the risk posed by a novel variant requires the degree of cross-reactive immunity between the new variant and previously circulating viruses to be assessed. In scenarios in which previous infections by antigenically related viruses have a limited effect, or promote infection with the novel variant, then the risk of the variant invading the host population successfully is substantially higher than in scenarios in which previous infections by related viruses are protective. Given the impact that different variants have had on transmission and control during the COVID-19 pandemic, fast detection and analysis of novel variants is essential for both national and global public health.

## Data Availability

All data produced are available online at https://github.com/yairdaon/waning

## Author Contributions

Conceptualisation: RNT, CPT, SI, UO; Methodology: RNT, UO, ES, FLR; Formal analysis: RNT, ES, YD, UO; Supervision: RNT, UO; Writing – Original draft: RNT, CPT, UO; Writing – Review and editing: All authors.

## Acknowledgments

Thanks to members of the Zeeman Institute for Systems Biology and Infectious Disease Epidemiology Research for useful discussions about this work.

## Data Availability Statement

Computing code developed to generate the results in the manuscript is available at https://github.com/yairdaon/waning

